# Bodily Self-Dysfunction in Psychosis: Altered Neural and Spinal Response to Self-produced Sensations

**DOI:** 10.1101/2025.01.28.25321269

**Authors:** Paula C. Salamone, Adam Enmalm, Reinoud Kaldewaij, Marie Åman, Charlotte Medley, Michal Pietrzak, Håkan Olausson, Andrea Johansson Capusan, Rebecca Boehme

**Author notes:** Authors contributed equally (Paula C. Salamone, Adam Enmalm).

## Abstract

Psychosis is often characterized by disturbances in the sense of self, with patients frequently misattributing self-produced sensations to external sources. While somatic hallucinations and misperceptions are common, the underlying disruptions in basic bodily self-processing remain unclear.

We aimed to investigate alterations in bodily self-processing, including touch and interoception, in psychosis using a multimodal, multi-method approach.

A total of 70 participants were included (35 patients diagnosed with psychotic disorders [DSM-IV F20-29], 35 age-and sex-matched controls).

Participants performed self-/other-touch-tasks and interoceptive assessments during functional magnetic resonance imaging (fMRI), evoked potentials (EP) measurements, and/or behavioral and psychophysical tests.

Primary outcomes included neural and behavioral responses to self-and externally-generated sensations (touch and heartbeat). Brain activation (fMRI), spinal responses (EPs), heartbeat perception and processing (EPs), and behavioral measures were analyzed, with preregistered hypotheses.

Patients demonstrated heightened neural activity during touch tasks, including increased right superior temporal gyrus activation during self-touch and heightened activity in a right temporoparietal cluster during social touch. Tactile self-other distinction impairments were evident at the spinal cord level (EPs). Behaviorally, patients showed reduced differentiation in tactile thresholds for self-vs. other-touch. Interoceptive impairments included diminished cortical responses to heartbeat signals (EPs), lower interoceptive accuracy (heartbeat detection), and reduced self-reported interoceptive sensitivity.

These findings reveal pervasive sensory and self-related disturbances in psychotic disorders. Impairments in differentiating self-and externally-evoked responses, detectable as early as the spinal cord level, may contribute to higher-order symptoms of psychosis.

## Introduction

Schizophrenia is often described as a disorder of the self^1,2^, involving heightened self-referential thinking^3^ and misidentifying self-produced sensations, such as perceiving one’s voice as alien^4^. The self is a hierarchical construct, with the bodily self forming its foundation^5,6^. A coherent bodily self-experience requires perceiving and identifying signals - proprioceptive, interoceptive, or somatosensory - as one’s own. Somatosensory signals are crucial for distinguishing self-evoked from non-self-evoked sensations, as self-touch and touch from others create the same peripheral stimulus^7^. This distinction is essential for self-functioning and interaction with the world.

We asked whether self-related symptoms in psychosis stem from a dysfunction in distinguishing self-from non-self-generated sensations. Using a multimethod approach, we assessed neural and behavioral responses to self-and non-self-generated sensations, exploring whether patients show alterations in self-related processes, at which level these occur, and their relation to symptomatology.

Previous studies have reported sensory domain alterations related to the bodily self in schizophrenia, yet these findings often lack integration across sensory modalities, and their neural underpinnings remain unclear. For instance, interoceptive impairments^8–10^, have been linked to positive symptoms^9,11^, though findings are inconsistent^10^ and may be confounded by cognitive factors. This can be addressed by a different task design paired with physiological measures like the heartbeat evoked potential (HEP). While altered HEPs in schizophrenia were reported, no behavioral measures were included^12^.

Tactile stimulation activates the somatosensory cortex, and social-affective touch also engages the insula^13^. Self-touch is processed differently^14^, likely due to its high predictability, leading to attenuated perception^15^. This attenuation is diminished in schizophrenia^16,17^, which might relate to positive symptoms^18^ and correlates with altered neural processing^19^. However, previous studies used tool-mediated setups, which, while controlled, lack ecological validity. Skin-to-skin touch offers a closed-loop for investigating sensory attenuation with greater relevance to real-world interactions.

The prevailing model of self-touch attenuation states that sensory outcomes are predicted, and when predictions match sensory outcomes, perception is attenuated^18^. We found differences between self-and non-self-touch already at the spinal cord level^20^ possibly due to modulation by top-down processes like predictions^21^. Sensory attenuation might be a factor mediating self-other-distinction: self-evoked sensations are always more predictable than externally evoked sensations. In line with this, the degree of neural self-other-touch distinction relates to higher order self-concept^14^, and the dissociative substance ketamine decreases this neural self-other-difference^22^.

Differences in sensory attenuation may reflect impaired predictive modeling in schizophrenia, for instance, it might result from weak low-level priors, causing imprecise predictions and prediction errors. This might be compensated for by strong higher order priors^23^, which then form the basis of delusional beliefs that explain away prediction errors^24^.

Here, we evaluated sensory distinctions between self-and externally generated sensations in touch and interoception. We used an ecological task with self-touch and other-touch during brain imaging, and during neurophysiology (evoked potentials) of brain and spinal cord. We measured sensory thresholds during self-touch and other-touch. We assessed interoception versus exteroception through behavior, electrophysiology, and self-report. The measures regarding interoception can be understood as mirroring the touch measures: they compare a self-generated (heartbeat) versus a non-self-generated (heartbeat-tone) stimulus.

Our preregistered hypothesis was to find altered distinction of self-and non-self-generated sensations (https://osf.io/kzscj/).

## Methods

### Procedure

The study was approved by the Swedish Ethics Authority. Participants provided written informed consent. An overview of the procedure is depicted in figure 1.

**Figure 1:**
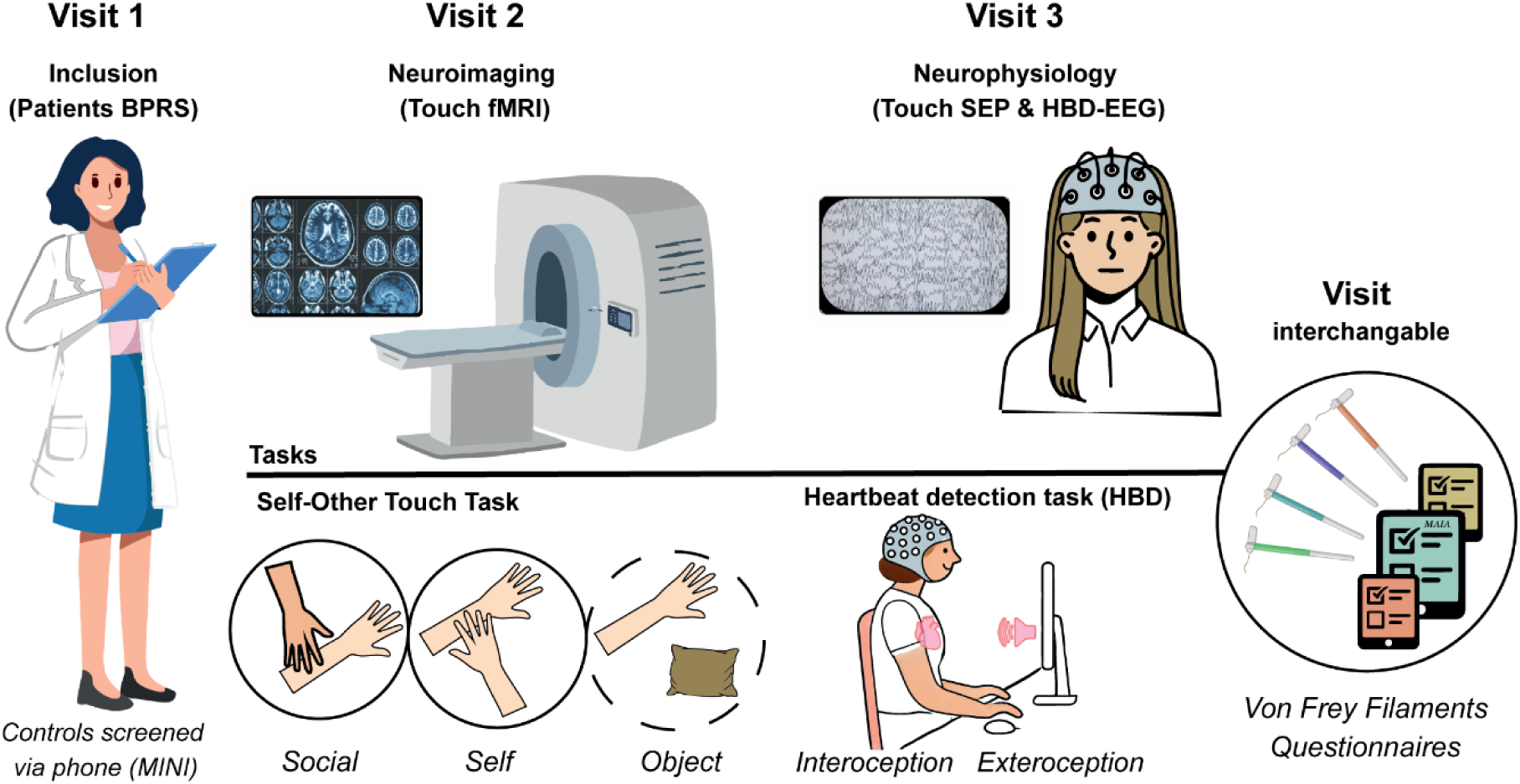
Procedure. Patients came for three visits, controls for two. The initial visit for patients included an interview using the Brief Psychiatric Rating scale (BPRS), while controls were screened over the phone using MINI-interview. fMRI = functional magnetic resonance imaging, SEP = somatosensory evoked potentials, HBD = heartbeat detection task, EEG = electroencephalogram

More details on all methods are provided in the supplement. If not state otherwise, analyses were preregistered (https://osf.io/e5gfu/).

### Participants

#### Patients

Recruited from outpatient psychiatric clinics in Östergötland, Sweden. Inclusion: psychotic disorder (DSM-IV, F20-F29), age 18-50. Exclusion: substance use (current/past 6 months), insufficient Swedish skills, intellectual disability, medical issues affecting left arm sensations.

#### Controls

Recruited via social media, flyers, and a participant database. Inclusion: no psychiatric disorders or major health issues, age 18-50. Exclusion: same as above.

#### fMRI

The touch paradigm consisted of three conditions, each presented 10 times in randomized order: self-touch, other-touch, and object touch for movement control. Participants performed or received slow, gentle strokes on their left forearm, or touched a pillow respectively. The protocol for image acquisition, scanner settings, and preprocessing was identical to our previous studies employing this task^14,22,25,26^.

Self-and other-touch were compared separately between groups using t-test. Results were corrected for multiple comparisons using the family-wise error (FWE) correction at whole brain level. In addition, we performed small volume FWE-correction (SVC) for a-priori regions of interest (ROI, anatomically defined): right posterior superior temporal gyrus (STG), insula, anterior cingulate cortex, primary somatosensory cortex, and precuneus. In an exploratory analysis, we included a functionally defined ROI of a temporoparietal cortex (TPC) region that showed significant effects in our previous pharmacological model of an altered self^22^.

#### Somatosensory Evoked Potentials (SEP)

As established previously^14,27^, SEPs were recorded during four conditions: baseline, self-touch, other-touch, and object-touch. Electrical stimulation targeted the radial nerve at the base of the left thumb, with 300 pulses delivered during two runs per condition.

Responses were recorded for 100ms after each pulse. Reponses were averaged across pulses and runs per condition. Analysis focused on amplitude and latency of the N13 (cervical) and N20 (cortical) components.

A repeated-measures ANOVA with the factors condition and group was used. An exploratory between-group Mann-Whitney-test on self-other difference was calculated.

#### Touch thresholds

As established previously^14,22^, touch thresholds were assessed using von Frey monofilaments (BIO-VF-M, Bioseb, Force of 0,008 g – 10 g / 0.078 mN - 98.066 mN) across) on the left dorsal forearm during four conditions: baseline (no touch), self-touch, other touch, and object touch. Thresholds were determined by the smallest filament detected in ≥5/10 applications.

A 2×2 ANOVA with factors group and condition was performed. An exploratory between-group Mann-Whitney-test on self-other difference was calculated.

### Interoception

#### Heartbeat detection

Electrocardiogram (ECG) was recorded. In the interoceptive condition, participants tapped a button upon perceiving their own heartbeat, with no external clue. The exteroceptive condition served as a control, where participants tapped in sync with a pre-recorded heartbeat sound (60bpm, irregular intervals). Each block lasted 2 minutes, with condition order counterbalanced. Accuracy was measured by tapping-cardiac synchronization (see supplement for accuracy and confidence score calculations)^28–30^. A 2×2 ANOVA was conducted, followed by post-hoc Tukey tests.

#### Heartbeat-evoked potentials (HEP)

Participants watched a monitor with a plus-sign. They were instructed to listen to a pre-recorded heartbeat (exteroception, as above), or to focus on feeling their own heartbeat (interoception) for 2min. The order was counterbalanced. EEG/ECG signals were recorded.

HEP was evaluated through combined EEG/ECG analysis. We compared overall HEP modulation, across conditions (interoception versus exteroception) and between groups using a point-by-point Monte Carlo permutation test with bootstrapping^31^.

#### Combined analyses

Combined analyses were conducted to explore the relationship between experimental measures and symptomatology (not pre-reregistered). Regressions predicted BPRS scores using group-differentiating experimental measures. Logistic regression assessed whether neural measures predicted group membership.

## Results

### Demographics and symptoms

60 patients expressed interest. 4 could not be reached, 11 withdrew interest after study description. Forty-five patients were scheduled for inclusion, 7 withdrew participation prior to inclusion, one did not meet inclusion criteria, and one withdrew after inclusion but prior to experimental procedures. 35 patients enrolled in the study (figure S1).

Diagnoses included schizophrenia (n=14), schizoaffective disorder (n=9), non-specific non-organic psychosis (n=8), delusional disorder (n=3), and acute schizophrenia-like psychosis (n=1). All patients were on antipsychotic medication (mean chlorpromazine equivalent 371.2±216.5mg). 19 patients were on stable antidepressants (see supplement). Patients had a mean illness duration of 4.8±4.5 years and mean BPRS score of 33.1±7.3.

85 controls expressed interest. 55 were eligible for participation. The 35 best matching for sex and age were included (figure S1). Group differences are summarized in table 1.

**Table 1:** Participant demographics and self-report outcomes.

| **Basic Demographics** | Controls | Patients | Test outcome | Significance |
| --- | --- | --- | --- | --- |
| Gender | 15F/20M | 15F/20M | $\chi^2 (1) = 0.0$ | $p = 1.000$ |
| Age | 35.7 (9.3) | 35.5 (7.4) | $U = 624.0$ | $p = 0.897$ |
| BMI | 24.3 (4.3) | 29.2 (6.1) | $t (68) = -3.882$ | $p < 0.001^{**}$ |
| **Handedness** | | | $\chi^2 (2) = 5.581$ | $p = 0.061$ |
| Right | 34 (97.1%) | 28 (80%) |  |  |
| Left | 0 (0%) | 4 (11.4%) |  |  |
| Ambidextrous | 1 (2.9%) | 3 (8.6%) |  |  |
| **Education** | | | $\chi^2 (2) = 7.921$ | $p = 0.019^{**}$ |
| Primary | 0 (0%) | 6 (17.1%) |  |  |
| Secondary | 13 (37.1%) | 15 (42.9%) |  |  |
| Tertiary | 22 (62.9%) | 14 (40%) |  |  |
| **Occupation** | | | $\chi^2 (2) = 22.2$ | $p < 0.001^{**}$ |
| Unemployed / sick | 0 (0%) | 18 (51.4%) |  |  |
| Student | 10 (28.6%) | 5 (14.3%) |  |  |
| Working | 25 (71.4%) | 12 (34.3%) |  |  |
| **Questionnaires** |  |  |  |  |
| MAIA | 101.5 (21.5) | 87.4 (18.3) | $t (68) = 2.952$ | $p = 0.004^*$ |
| MAIA: Noticing | 3.6 (0.8) | 3.2 (0.9) | $t (68) = 1.964$ | $p = 0.054$ |
| Maia: Non-distracting | 2.4 (0.8) | 2.1 (0.8) | $t (68) = 1.787$ | $p = 0.078$ |
| MAIA: Non-worrying | 3.1 (0.9) | 2.3 (1.2) | $t (68) = 2.941$ | $p = 0.004^*$ |
| MAIA: Attention regulation | 3.2 (0.9) | 2.9 (0.9) | $t (68) = 1.243$ | $p = 0.218$ |
| MAIA: Emotional awareness | 3.4 (0.9) | 2.9 (1.1) | $t (68) = 2.024$ | $p = 0.047$ |
| MAIA: Self-regulation | 3.0 (1.0) | 2.5 (1.1) | $t (68) = 1.927$ | $p = 0.058$ |
| MAIA: Bodily listening | 2.5 (1.1) | 2.5 (1.2) | $t (68) = 0.244$ | $p = 0.808$ |
| MAIA: Trusting | 3.8 (1.0) | 3.0 (1.4) | $U = 823.5$ | $p = 0.013$ |
| STQ | 37.2 (9.1) | 44.1 (14.0) | $U = 442.5$ | $p = 0.046^*$ |
| EQ | 46.1 (13.0) | 41.4 (12.8) | $t (68) = 1.510$ | $p = 0.136$ |
| SP: Low Registration | 28.5 (5.6) | 33.9 (7.5) | $U = 335.0$ | $p = 0.001^*$ |
| SP: Sensation Seeking | 42.6 (5.9) | 41.1 (7.5) | $t (68) = 0.905$ | $p = 0.369$ |
| SP: Sensation Sensitivity | 33.1 (5.6) | 37.9 (9.5) | $U = 406.5$ | $p = 0.016$ |
| SP: Sensation Avoiding | 36.1 (7.1) | 40.7 (9.1) | t (68) = -<br>2.401 | p = 0.019 |
| SP: Touch processing | 28.6 (4.1) | 30.4 (6.6) | U = 522.5 | p = 0.292 |

*Note. Demographics and questionnaire results. The Multidimensional assessment of interoceptive awareness (MAIA) subscales and Sensory profile (SP) subscales were corrected for multiple comparisons using Bonferroni-Holm correction. \*Significant difference. STQ = Social Touch Questionnaire; EQ = Emotional quotient.*

### Increased neural responses to touch in patients

During self-touch, patients exhibited significantly higher activation in the right STG (*p*_FWE(SVC)_=0.028, MNI_xyz_=48,-26,14; *p*_FWE(SVC)_=0.042, MNI_xyz_=42,-30,16, Figure 2A), potentially reflecting altered predictive coding.

**Figure 2:**
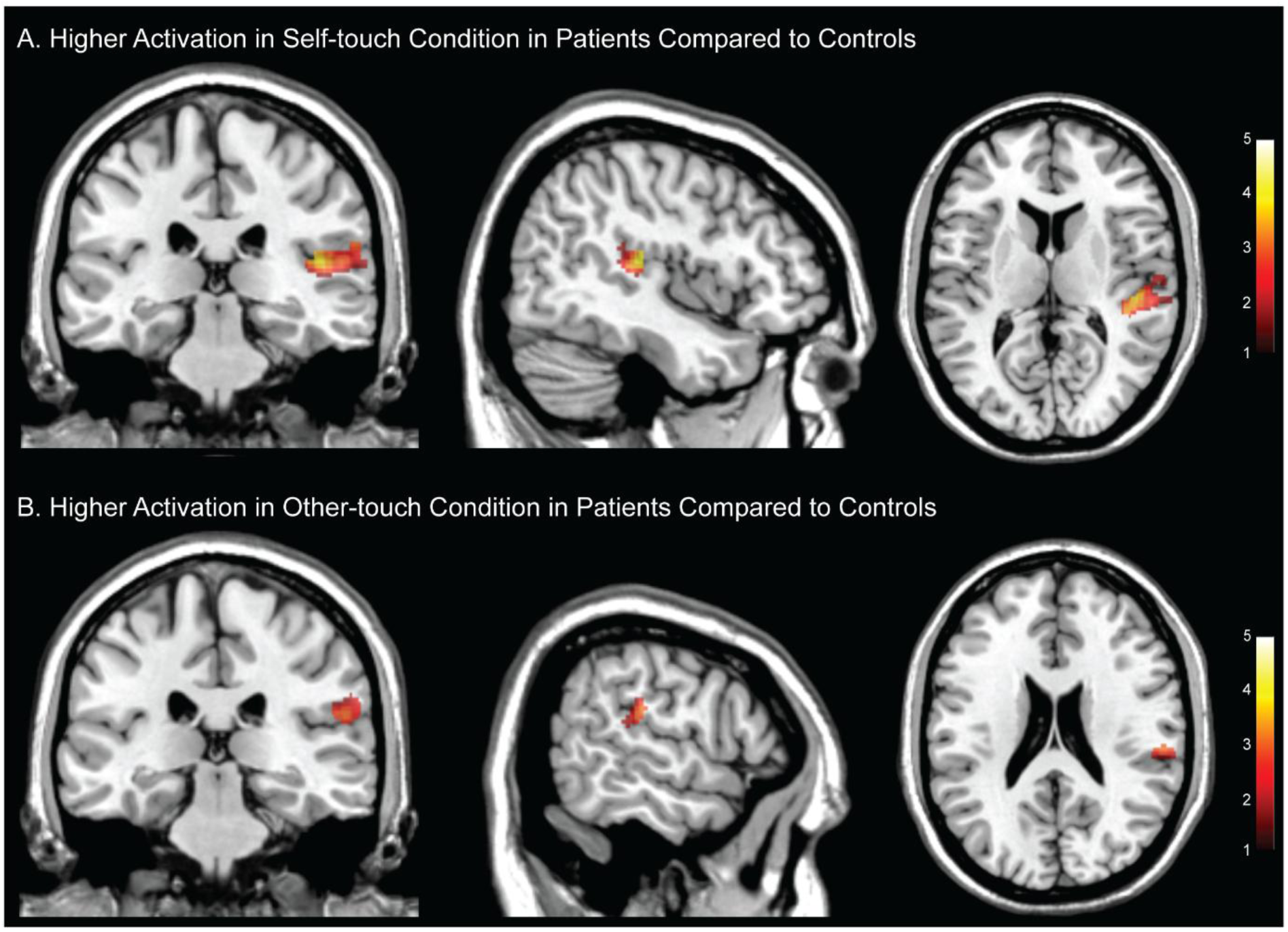
Brain activity differences. The patient group compared to the controls showed increased activity increased during (A) self-touch in the superior temporal gyrus (MNI_xyz_ = 45, - 28, 11) and during (B) other-touch in the temporo-parietal cortex (MNI_xyz_ = 59,-28, 22). Thresholded at p<0.001 for display purpose, color-bar indicates t-values.

Exploratory analysis of other-touch revealed greater activation in the TPC in patients (p_FWE(SVC)_=0.049,MNI_xyz_=56,-26,20, Figure 2B).

### Reduced latency difference between self-and other-touch at the spinal cord level

In the spinal cord, there was a significant condition x group interaction (F(2,128)=3.119, p=0.048) on N13 latency. Follow-up group comparison revealed significantly smaller latency differences between self and other in patients (U=793, p=0.018, Figure 3A), suggesting impairments in early sensory processing.

**Figure 3:**
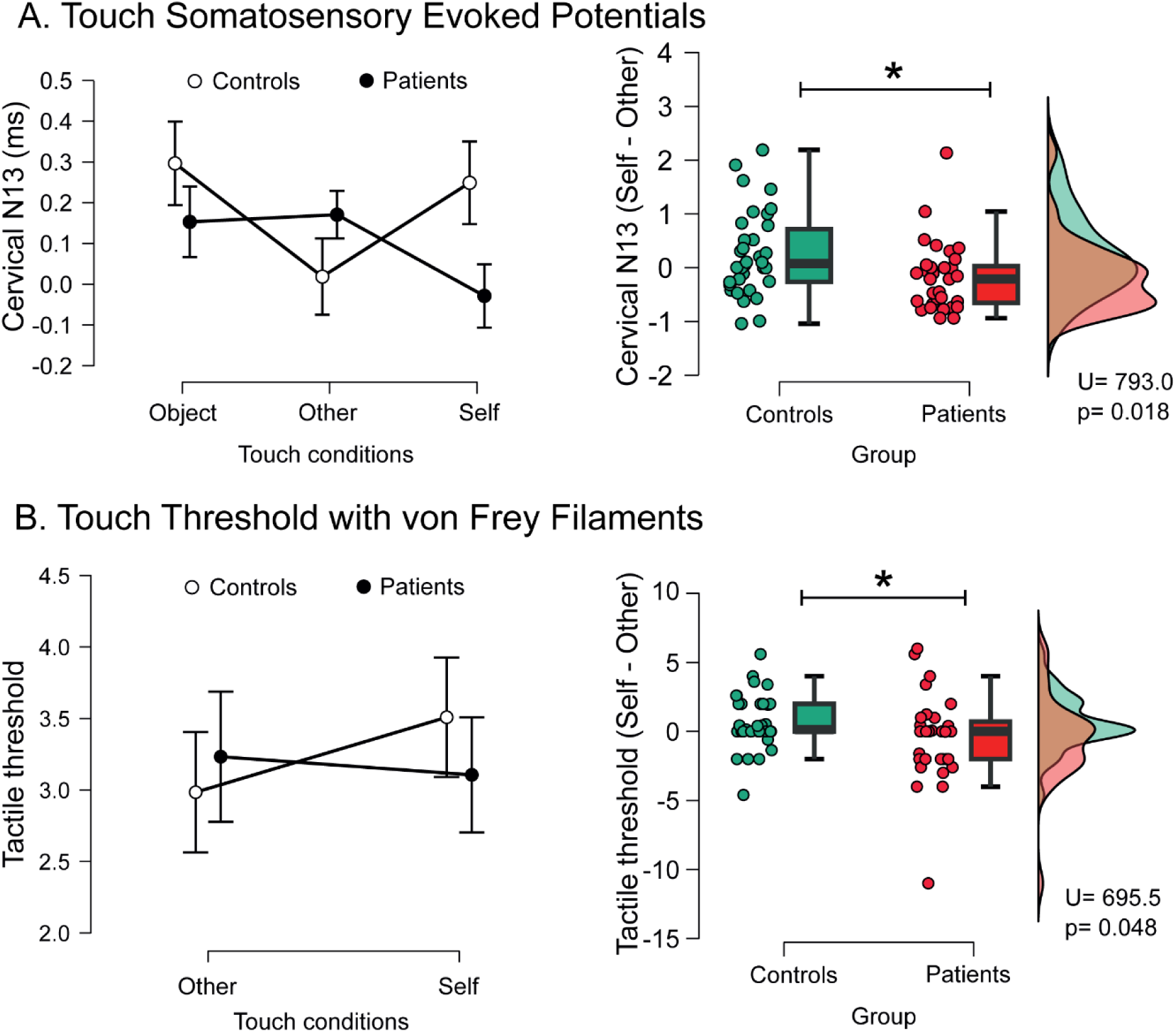
Self-other touch task during somatosensory evoked potentials (SEPs) and touch detection thresholds. (A) Spinal cord SEP (N13): Significant condition and group differences in N13 latency (left). Self-other difference was lower in patient (right). (B) Touch detection thresholds: No main effect of condition or group (left). Self-other difference in thresholds was lower in patients compared to controls.

### Reduced difference between detection thresholds during self-other-touch

An exploratory analysis revealed a significant group effect for threshold differences during self-and other-touch (U=695, p=0.048, Figure 3B). Unlike controls, who exhibited higher thresholds during self-touch, patients showed comparable thresholds across conditions, indicating altered sensory attenuation mechanisms.

### Reduced interoceptive accuracy

Patients exhibited reduced interoceptive accuracy (main effect of group: *F*(1,127)=6.430, p=0.012, group x condition: *F*(1,127)=4.051, p=0.046). A post hoc test confirmed a significant group difference in the interoceptive condition (t=-3,180, p=0.01, Figure 4D), where patients displayed lower accuracy.

**Figure 4:**
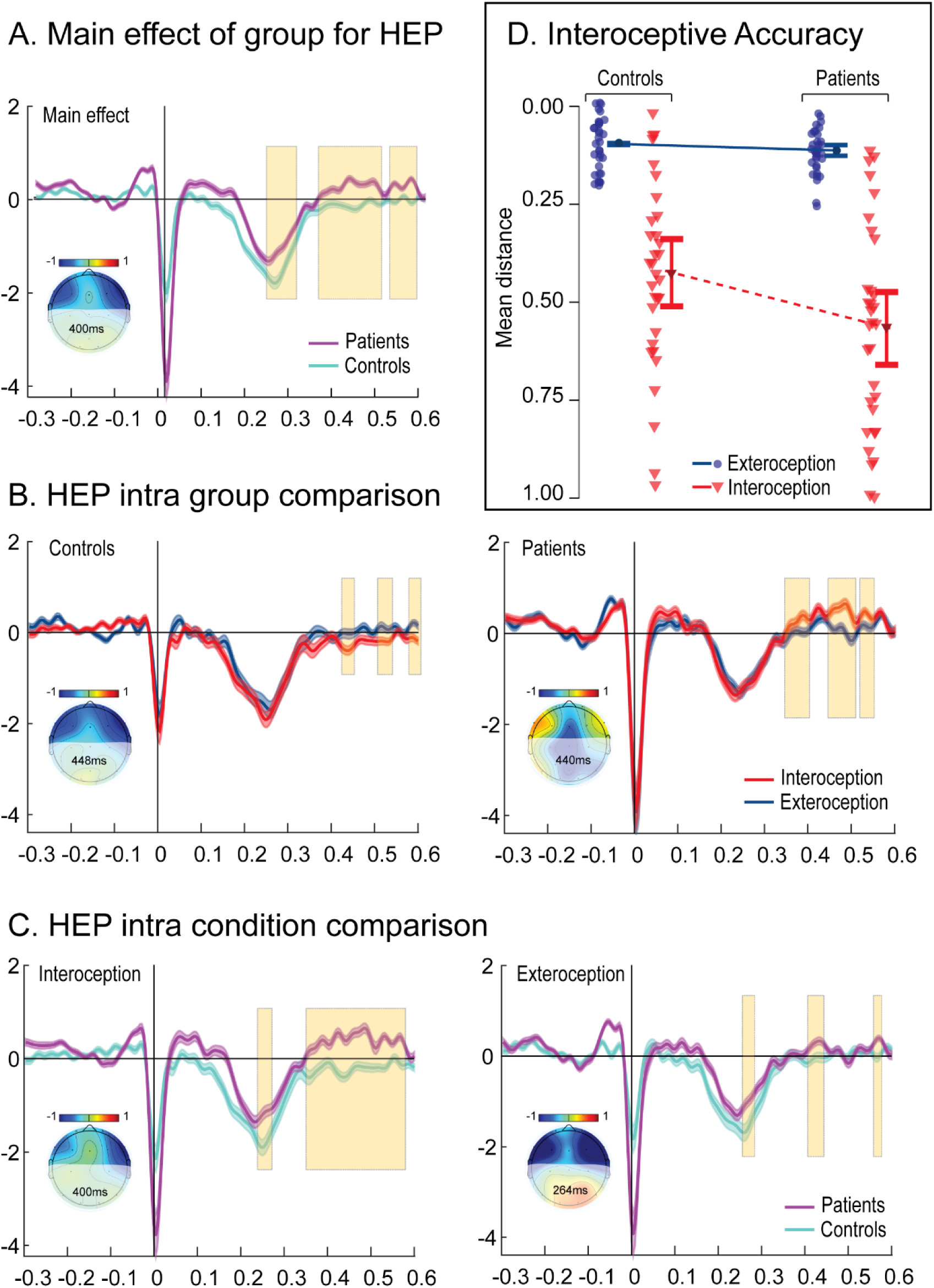
Heartbeat Detection Task, Cortical and Behavioral Measurements. (A) Main effect of group for HEP: Comparing controls (teal) and patients (purple). (B) HEP intra-group comparison: Controls (left) and patients (right). Interoception (red) vs. exteroception (blue). (C) HEP intra-condition comparison: Interoception (left) and exteroception (right). Patients (purple) vs. controls (teal). Mean modulations shown with standard error of the mean (SEM, shadowed lines). Significant time windows marked with yellow shading. Topographical maps highlight activity differences at indicated time points. (D) Interoceptive accuracy: Mean distance indices for patients (red) and controls (blue). Interoception (triangles) vs. exteroception (circles) show significant main effect of group and condition, and interaction effects. Controls had higher accuracy than patients in interoception. No group differences in exteroception. Both groups had better exteroception accuracy versus interoception. Error bars represent SEM.

### Altered interoceptive HEP modulation

Patients showed lower HEP amplitudes across three time windows (236-30ms, 356-500 ms, 520-580ms, Figure 4A). Patients did modulate HEP differently for intero-and exteroception, but showed the opposite pattern of controls, i.e. modulation was higher during exteroception than interoception(Figure 4B).

For interoception, patients showed lower HEP modulation over a longer period (240-272ms, 352-580ms) than during exteroception (256-284ms, 408-444ms, Figure 4C).

### Relation to symptomatology and group membership

Touch-related measures significantly predicted the BPRS total scores (F(4,24)=3.26, p=0.033, adjusted R²=0.274), with touch threshold difference between self-and other-touch showing the strongest predictive value (t=-2.14, p=0.045).

Neural measures significantly predicted group membership (X^2^=17.9, p=0.001) with STG activation during self-touch as the strongest predictor (Wald statistic=7.4, p=0.007, Odds Ratio 52.8, Table S6). The model explained 39.5% of the variance (Nagelkerke R^2^) and predicted 66.7% of the cases correctly.

## Discussion

Patients with psychotic disorders displayed reduced neural attenuation of self-evoked sensations, and a smaller difference between self-and non-self-evoked signals already at the spinal cord level. This was accompanied by smaller differences in perceptual thresholds during self-vs. non-self-evoked touch. In the second sensory domain, interoception, patients displayed lower accuracy in detecting the signals originating from self (i.e. heartbeats) and a dysfunctional modulation of the neural markers of these signals (HEP). This was especially pronounced during interoceptive focus. These alterations align with predictive coding models of psychosis, suggesting a shared mechanism underpinning deficits in self-referential processing. Touch-related variables were also associated with symptom severity, and neural measures reliably predicted group membership.

We did not observe the expected smaller self-other distinction in touch-related brain activity. Instead, both touch conditions showed increased activity. Patients exhibited specific alterations: elevated neural activity in the right STG during self-touch and in a right temporoparietal cluster during other-touch. These differences may stem from early-stage or low-level touch processing dysfunction, potentially at the spinal cord level, as suggested by neurophysiological findings of reduced latency differences between self-and other-touch at the cervical spine, driven by faster self-touch latencies.

In interoception, patients showed multidimensional alterations: reduced cortical interoceptive modulation (HEP amplitude), decreased interoceptive accuracy (HBD task), and lower sensitivity (MAIA), aligning with our hypotheses and previous findings.

### Touch

Our findings of reduced attenuation of self-evoked sensations in psychosis extend previous research^18,19^, demonstrating these deficits across both cortical and spinal levels. Altered spinal processing, reflected by reduced latency differences between self-and other-touch, highlights dysfunction in early-stage sensory processing, potentially originating at the dorsal horn^32^. This novel observation suggests that sensory abnormalities in psychosis may begin at subcortical levels and influence higher-order processes.

Previous studies reported also an overall reduced neural response to touch^19^, while we found overall increased response. This difference may stem from their use of mechanical tapping, engaging different pathways than our affective touch stimulus^13^. The nature of touch (social vs. non-social) likely influences patients’ general response but diminished self-touch attenuation appears consistent across touch types.

During self-touch, patients showed increased STG activation, which significantly predicted group membership. In contrast, healthy populations typically show STG deactivation during self-touch^14,26^. Increased STG activation suggests a mismatch between sensory predictions and incoming signals, potentially driven by weak priors or disrupted top-down modulation. The superior temporal areas are involved in various functions, including speech processing, biological motion^33^, and social interaction^34^. STG abnormalities are implicated in hallucinations and self-other distinction^35–37^, supporting this region’s role in psychotic symptomatology. A potential common underlying mechanism might be related to predictive processing^38^. These findings have led to suggestions of STG as a target for neurofeedback intervention to reduce self-symptomatology^39^. Therefore, the here reported increased STG response to self-touch in patients may result from a mismatch between prediction and sensation, potentially due to faulty predictions^40^, altered bottom-up sensation, or a failure to integrate sensory evidence and update predictions^41^.

Altered SEP latencies suggest a breakdown in self-other distinction at the spinal level, potentially driven by disruptions in predictive modulation from cortical circuits. Few studies have examined cortical SEPs in this group, with mixed results^42–46^. Our SEP design might be more sensitive as it integrates evoked potentials with additional touch stimuli and applies baseline correction removing bias by factors like height and nerve conduction speed.

In controls, simultaneous other-touch reduces SEP latency in both the spinal cord and brain compared to self-touch^14^, indicating potential top-down modulation. This effect was replicated in controls but absent in patients, suggesting sensory pathway alterations at the dorsal horn. Similar spinal cord-level changes have been linked to increased somatosensory sensitivity in an autism animal model^47^, and at least one gene (GABRB3) implicated in this model is associated with schizophrenia risk^48^. These findings align with reports of generalized sensory abnormalities and higher touch aversiveness in psychosis patients^49^.

Cortical dysfunction may influence spinal cord alterations, as somatosensory circuits in the spinal cord receive significant input from cortical neurons^32^. Altered predictive processing in schizophrenia could impact tactile signals at the spinal cord level, aligning with the predictive coding model of psychosis^50^ and our data showing differential spinal processing during self-and other-touch in controls^14^.

Studies on audition and vision in schizophrenia report altered evoked potentials, with reduced gating in paired-pulse designs^51,52^, reflecting difficulties attenuating irrelevant stimuli. This aligns with our findings of increased activity and shorter latency during self-touch, a highly predictable and usually irrelevant stimulus for controls.

To test basic tactile sensitivity, we measured detection thresholds and found no baseline differences between patients and controls but a significantly smaller threshold difference between self-and other-touch in patients. This supports altered integration of additional stimuli during self-and other-touch, consistent with our SEP findings and overall results.

Our findings underscore the importance of targeting sensory dysfunction therapeutically, potentially through sensory training or pharmacological modulation of spinal circuits.

### Interoception

Patients exhibited interoceptive processing alterations both behaviorally and neurally, which aligns with a disrupted bodily self-representation in psychosis. Patients showed reduced interoceptive accuracy (i.e. specific to self-generated stimuli), with no impairments in exteroceptive trials, aligning with previous research^9,10^. The lack of group differences in exteroceptive conditions suggests the results were not due to cognitive or motor impairments. Reduced HEP modulation further supports this, as HEP reflects cortical processing of interoceptive signals integral to maintaining allostasis and self-awareness. The overall directionality is consistent with resting-state HEP-findings^12^. Monitoring one’s heartbeat is crucial for regulating allostasis, brain activity, responsivity, and the bodily-self model^53,54^. Reduced HEP modulation, linked to less accurate bodily signal perception, may reflect or contribute to a faulty bodily-self model, as larger HEP modulations relate to self-referential thoughts^55^ and are diminished in depersonalization^56^. The reversal of modulation patterns in patients suggests a maladaptive predictive coding mechanism, where interoceptive priors may dominate over sensory evidence. Such dysfunctions could underpin depersonalization and impaired self-referential processing in psychosis.

### General discussion

Psychosis patients showed altered processing of self-originating stimuli across domains. Following predictive processing theories in schizophrenia^24,50^, the increased neural response to self-touch may result from faulty predictive coding, such as weak low-level priors or failure to update predictions due to strong high-level priors. This mechanism could also explain reduced interoceptive accuracy. Aberrant sensations from such dysfunctions may underlie psychotic symptoms, as suggested by the aberrant salience hypothesis^57^ and patients’ prodromal experiences^58^.

Touch-related measures predicted symptomatology, particularly in the negative and affective domains. This differs from prior findings linking reduced attenuation to positive symptoms, possibly due to our medicated, chronic-phase sample with low positive symptoms. Interoceptive self-models likely rely on multiple interoceptive sensations, not heartbeat alone^59^. Touch, bridging extero-and interoception^60^, is central to predictive models of self and world. The STG’s increased activation during self-touch and altered spinal cord self-touch processing further highlight the importance of self-and touch-related processes in schizophrenia.

### Limitations

Our sample showed low symptomatology, was medicated, and had the condition for several years. Self-reported sensory sensitivity and avoidance were higher in the patient group, which might affect social touch processing. We included patients of any psychotic disorders (DSM F20-29), which may introduce variability in findings but enhances generalizability. Previous studies have found only limited neural differences between schizophrenia and schizoaffective disorders^61^.

## Conclusion

Psychosis patients display significant dysfunction in the processing and perception of self-evoked sensations, across touch and interoception, evident at cortical and subcortical levels. We suggest a dysfunctional bodily-self predictive model as the shared underlying mechanism.

Our findings support predictive coding theories, where disrupted priors or impaired sensory integration contribute to aberrant self-referential processing in psychosis. Reduced attenuation of self-generated signals, across touch and interoception, may lead to altered salience attribution, a hallmark of psychosis. Future research should explore whether interventions that enhance sensory integration or recalibrate predictive coding can ameliorate symptoms.

## Data Availability

All data produced in the present study are available upon reasonable request to the authors.

## Acknowledgements

This study was funded by grants from the Swedish Research Council (2019-01873, 2023-02116), ALF Grants, Region Östergötland (RÖ-960561, RÖ-1000522), grants from Medical Research Council of Southeast Sweden (FORSS-940816, FORSS-969464), and Åke Wiberg stiftelse (M19-0369) awarded to RB, and Fredrik och Ingrid Thurings Stiftelse (2021-00688) to PS.

