## Supplemental Material for "Bodily Self-Dysfunction in Psychosis: Altered Neural and Spinal Response to Self-produced Sensations"

*Authors contributed equally

**List of included elements:**

1. Supplementary text: methods, results, discussion
2. Supplementary tables:
   a. S1-4: Demographics of the subsamples per task
   b. S5: Main group effect of fMRI results
   c. S6: Logistic regression analysis
3. Supplementary figures:
   a. S1: Flow chart of participant attrition
   b. S2: Main effect of group, brain activity difference for patients > controls
4. Supplementary references

**Methods:**

Procedure

Patients attended three visits, controls two. At visit 1 (only patients), patients were interviewed by a trained psychiatrist using the Brief Psychiatric Rating Scale (BPRS) to assess psychotic symptoms. Controls were interviewed on the phone by a trained staff member using the MINI questionnaire. At Visit 2 (patients and controls), participants underwent neuroimaging at the Center for Medical Imaging and Visualization in Linköping University Hospital, where they performed the self-other touch task. Prior, all participants were instructed to perform the touch inside a dummy scanner. At Visit 3 (patients and controls), participants underwent a neurophysiological evaluation at the Neurophysiology Clinic in Linköping University Hospital. Participants performed the self-other touch task while somatosensory evoked potentials were recorded from the somatosensory cortex and the spinal cord at C6. Additionally, they performed a heartbeat detection task to evaluate interoception at the Center for Social and Affective Neuroscience during EEG recordings. The order of visits 2 and 3 was interchangeable, each lasting 1-2 hours. During one of the visits, touch detection thresholds during the self-other touch task were measured and questionnaires were filled in. Not all participants finished each task (figure S1, tables S1-4). During data analyses, outliers >2.5SD from the mean were excluded.

Chlorpromazine equivalents

Current medication was extracted from medical records and chlorpromazine equivalents were calculated in following^1^. In cases where the long-acting injectable (LAI) used by the patient was not included in the conversion tables developed^1^ the LAI was converted to oral form in accordance with tables provided by The American Psychiatric Association^2^ before conversion to chlorpromazine equivalents.

Antidepressant medication

A total of 19 patients were medicated with antidepressant medication. Fluoxetine equivalents were calculated for antidepressant medication^3^. Three patients were medicated with Duloxetine which was not converted into Fluoxetine equivalents. One of these patients was medicated with both Duloxetine and Mirtazapin, and only had the Mirtazapin medication converted and considered for further analysis. Mean Fluoxetine equivalent dose of the 17 considered patients was 31.4 mg (range: 11.1-51.9 mg).

Touch task fMRI

Through a pair of MR-compatible goggles (VisuaStim Digital; Resonance Technologies), participants were prompted on the upcoming condition. The instructions were in Swedish: 1) “Active, please stroke your arm”; 2) “Passive, the experimenter will touch your arm”; and 3) “Active, please stroke the object”. Participants were shown instructions with white text for three seconds (cue phase). After three seconds, the text switched to green, signaling that touch should be initiated (touch-phase). The touching continued for 12 seconds, after which the text was replaced with a plus sign and the touching ended. A 12 second rest period followed. The experimenter was always a woman, standing next to the scanner where she received auditory cues on when to perform the other-touch condition. Each condition was repeated 10 times in a random order, for a total task duration of approximately 13.5 minutes.

Data acquisition fMRI

Functional magnetic resonance imaging (fMRI) data was acquired by a 3.0 Tesla scanner (Prisma, Siemens) with a 64-channel head coil. The following settings were used: for T1-weighted anatomical images: repetition time = 2300ms; echo time = 2.36s; flip angle = 8°; Field of view = 288 x 250 mm^2^; Voxel resolution = 0.97 x 0.87 x 0.90 mm^3^, for echo planar images (EPI): repetition time = 1030 ms; echo time = 30 ms; slice thickness = 3 mm; matrix size = 64 x 64; field of view = 192 x 192 mm^2^; in-plane voxel resolution = 3 mm^2^; flip angle = 63°).

MRI data were preprocessed and analyzed in Matlab (MathWorks, MA, USA) using SPM12 (SPM, Wellcome Department of Imaging Neuroscience, UK; http://www.fil.ion.ucl.ac.uk/spm). The preprocessing consisted of motion correction; co-registration of the mean EPI and anatomical (T1) image; segmentation of T1 image and spatial normalization to Montreal Neurological Institute (MNI) template; application of normalization parameters to all EPI volumes; spatial smoothing with an isotropic Gaussian kernel of 6-mm full width at half-maximum.

The first level model included the cue-phase, the touch-phase, a regressor for touch offset one second after each active condition, and realignment parameters. In addition, the first temporal derivative of realignment parameters in x,y,z-directions and a regressor censoring volumes with *>*1 mm volume-to-volume movement were added to the model, to account for the potential of increased movement.

To compare self-other-distinction between groups, we calculated a contrast between self-touch (motion corrected by using self>object-touch contrast) and other-touch at the first level. At the second level a t-test compared groups. To assess the differences between groups independent of condition, a flexible factorial ANOVA with the factors group and condition was used. To compare the conditions separately, between-group t-test was used.

SEP data acquisition

Data were acquired using the NicoletEDX system with an AT2+6 amplifier (Carefusion) and recorded via Synergy 20.0 (Carefusion). Following the standard clinical protocol, the pulses were individually adjusted to induce a small twitch in the thumb yet remain at a tolerable level. The average intensity of stimulation was 11.0 +- 2.6 (Range 5.9-17.3). Recording electrodes were placed at the C6 cervical level, and the C4, CZ, and FZ scalp positions. Skin impedance at the electrode sites was maintained below 8 kΩ. Recordings were referenced online to Fz, bandpass-filtered from 2 Hz to 2 kHz, with an amplifier range of 5 mV and display sensitivity of 20 μV per division.

Baseline-to-peak amplitudes were automatically calculated, with the baseline defined as the point immediately before the average waveform. Peaks were visually inspected and, in case the automatic detection was incorrect, adjusted manually. Data of the three touch conditions were corrected using the baseline condition to control for height and nerve conductance speed.

Heartbeat detection task (HBD) analyses

We deviated from the preregistered analysis plan to calculate a metacognition index based on the difference between objective accuracy and subjective confidence. We changed to the accuracy index because it is based on a newer, less biased method that calculates synchronization accuracy and is uniquely scaled. This index represents accuracy as a mean distance, with zero indicating optimal performance. Increased distance indicates decreased accuracy, complicating a direct comparison between maximum mean distance and minimum confidence scores. This synchronization index reflects the participant's ability to adjust responses to cardiac rhythms without bias from the total number of responses. Therefore, we followed a similar analysis approach, conducting a 2x2 ANOVA and post hoc t-tests with a Tukey correction.

HBD subjective ratings

At the end of each block, participants were asked whether they were sensing their heartbeat and to rate their confidence in their performance across both conditions.

HEP data acquisition

EEG and ECG recordings were acquired using a BIOPAC B-Alert 24-channel system, which comprises 20 active EEG channels, 2 mastoid references, and 2 chest-mounted ECG channels. Data were collected at a sampling rate of 2000 Hz with Acqknowledge software (Biopac) and underwent standard preprocessing as outlined in previous studies^4^.

HEP data processing

The EEG signals were downsampled offline to 250 Hz and bandpass-filtered between 0.5 - 30 μV. The HEP-lab toolbox in Matlab^5^ was used to align ECG events with the continuous EEG signal. To address artifacts related to eye movements, blinks, and cardiac activity, independent component analysis (ICA) and a visual inspection protocol were implemented, based on established methods^6,7^. Any noisy electrodes were excluded from ICA and later interpolated. As early time windows may still carry cardiac field artifacts, only data from 200 to 600 ms post-R-peak from the ECG were analyzed^7,8^. EEG data were divided into epochs spanning -300 to 600 ms and baseline-corrected from -300 to 0 ms relative to the R-peak. Analysis was conducted within a fronto-central region of interest (ROI) including channels Fp1, Fp2, F7, F3, Fz, F4, and F8.

To control for potential confounders, we extracted HEP modulation from the time windows identified as exhibiting significant differences during our initial analysis (main effect of HEP at the group level). This extraction involved calculating the negative area under the curve (AUC) for the mean HEP across the specified time windows (time-window 1: 236-304 ms, time-window 2: 356-500 ms, time-window 3: 520-580 ms) for each participant. We subsequently performed a two-way ANOVA between group and time, revealing a significant effect for group (F(1, 180) = 4.666, p = 0.032) and time (F(2, 180) = 18.574, p < 0.001). Following this, we correlated the average AUC HEP index with potential confounding factors that showed significant differences between groups, specifically medication, body mass index (BMI), and heart rate (HR).

We performed an exploratory analysis to assess HR and heart rate variability (HRV) across groups and conditions with a repeated measures ANOVA. The aim of this analysis was to explore if there was a group by condition interaction that could bias results. ECG data was transformed in Matlab to calculate the R-R interbeat interval that was later analyzed with Kubios software^9^. This is an automated HRV analysis tool for both time and frequency domains, optimized for short intervals. Using an autoregressive algorithm, Kubios computed the power spectrum, distinguishing among high frequency, low frequency , and very low frequency bands. The low/high ratio, a common marker of sympatho/vagal balance, served as an HRV index. For consistency, frequency components were calculated in normalized units (n.u.), representing each component's power relative to total power minus the very low frequency bands.

Combined analyses

To understand whether measures related to symptomatology, we performed three linear regressions on BPRS scores (1) with the experimental measures that differed between groups, (2) with touch-related measures, and (3) with interoception-related measures. The measures included beta-values from fMRI ROIs (STG during self-touch, TPC during other-touch), SEP latency self-other-difference, threshold self-other-difference, HEP AUC during interoception, HBD accuracy difference between intero- and exteroception. Self-reports were not included.

We tried to predict group membership from neural measures, using a logistic regression with the regressors: STG ROI during self-touch, TPC ROI during other-touch, SEP latency self-other-difference, HEP AUC during interoception.

To understand the relationship between neural measures of the touch and interoception modalities, we used a linear regression on HEP AUC during interoception with the touch-measures as predictors (beta-values from the STG ROI during self-touch, beta-values from the TPC ROI during other-touch, SEP latency difference between self- and other-condition) and group as a factor.

**Results**

fMRI

*Main Group Differences in Self-Other Touch Processing*

Across both conditions, (i.e. the main group effect), patients showed higher activation compared to controls, primarily in sensory and associative brain regions, including the occipital cortex, middle temporal gyrus, fusiform gyrus, postcentral gyrus, precentral gyrus, supramarginal gyrus, and superior parietal lobe (Table S5, Figure S2). Controls exhibited greater activation in the right occipital cortex, right calcarine cortex, and right cuneus compared to patients. In contrast to our pre-registered hypothesis, the group comparison for the difference between self- and other-touch processing did not reveal significant group effects.

There were no significant effects for the ROIs other than the ones reported in the main manuscript.

SEPs

*Spinal cord*

There was no significant main effect of condition (𝐹(2,128)= 1.305, 𝑝=0.275) or group F(1,64)= 0.323, p= 0.572) on N13 latencies.

Regarding amplitudes in the spinal cord (cervical C6), we found no main effect of condition (𝐹(2,130)= 0.968 𝑝=0.383) or group F(1,65)= 0.299, p= 0.587) and the interaction was not significant (F(2,128)= 2.506, p=0.086).

*Cortex*

At the cortical level, we found no significant differences at Cz or C4 regarding the N20 latencies. The results for Cz were: no main effect of condition (𝐹(2,132)= 0.886, 𝑝= 0.415), group F(1,66)= 0.509, p= 0.478), or interaction (F(2,132)= 1.676, p=0.191). The results for C4 were: no main effect of condition (𝐹(2,134)= 0.380, 𝑝= 0.685), group F(1,67)= 0.530, p= 0.469), or interaction (F(2,134)= 0.634, p=0.532).

Amplitude of the SEP at the cortical level showed similar effects for both Cz and C4 electrodes. Cz amplitudes showed a main effect of condition (𝐹(2,126)= 13.502, 𝑝< 0.001), but no effect of group 𝐹(1,63)= 0.142, p= 0.708) and no interaction (𝐹(2,126)= 0.925, 𝑝= 0.399). Similarly, C4 amplitudes showed differences for condition (𝐹(2,130)= 11.762, 𝑝< 0.001), but no effect of group 𝐹(1,65)< 0.001, p= 0.988) and no interaction (𝐹(2,130)= 0.230, 𝑝= 0.795).

The exploratory self-other-difference comparison between groups showed no significant difference for either Cz (t= 0.965, 𝑝= 0.338) or C4 (U= 629.500, 𝑝= 0.683).

Control analysis of SEP stimulation intensity

An independent samples t-test was conducted on stimulation intensity for the SEP. No significant difference between groups were found (t(68)=0.938, p=0.351).

Touch detection thresholds

There were no significant results for condition (𝐹(2,128)= 0.218, 𝑝= 0.642), group 𝐹(1, 128)= 0.033, 𝑝= 0.856), or interaction 𝐹(1, 128)= 0.584, 𝑝= 0.446).

HBD – exteroception accuracy and subjective ratings

There were no significant differences between groups for the exteroceptive condition (t=-0.374, p= 0.982). Both groups had higher accuracy during exteroception compared to interoception (controls: t= -7.462, p< 0.001; patients: t= -10.244, p< 0.001).

Confidence score analysis revealed a main effect of condition (𝐹(1,132)= 87.039, 𝑝< 0.001), but there was no effect of group (𝐹(1,132)= 1.776, 𝑝< 0.185), or interaction (𝐹(1,132)= 0.197, 𝑝= 0.658). Posthoc assessment indicates that both controls (t=6.911, p< 0.001) and patients (t= 6.283, p< 0.001) rated confidence higher for exteroception, as expected for a clearly perceivable stimulus.

Similar results were found for the detection of the signals: there was a main effect of condition (𝐹(1,132)= 99.029, 𝑝< 0.001), but there was no effect of group (𝐹(1,132)= 1.283, 𝑝< 0.259), or interaction (𝐹(1,132)= 0.519, 𝑝= 0.519). Posthoc assessment indicates that both controls (t=7.494, p< 0.001) and patients (t= 6.579, p< 0.001) detected the exteroceptive signals more clearly.

HEP

Within groups, we found that the controls displayed the expected HEP modulation, with higher amplitudes during the interoceptive than during the exteroceptive condition in three time-windows (time-window 1: 424-452 ms, time-window 2: 508-540 ms, time-window 3: 580-600 ms, Figure 4B).

*Control analysis of HEP*

The results in patients were not associated with their medication intake (rho= -0.033, p= 0.864). Additionally, HEP modulation in the total sample was not associated with BMI (rho= 0.082, p= 0.531). HEP was associated with HR (rho= 0.459, p< 0.001), but not HRV (rho= 0.003, p= 0.982). Although HR can influence overall HEP, we found similar correlation for both groups (C: rho= 0.407, p= 0.023, P: rho= 0.463, p= 0.012). Therefore, HR might be related to HEP, but it does not explain the group differences.

*Heartrate*

We found that there was a main effect of condition (F (1, 63)= 8.610, p= 0.005) and group (F (1, 63)= 5.586, p= 0.021). Both groups displayed a lower HR during interoception (patients mean= 81.120, SD= 13.672; controls mean= 73.182, SD= 12.670) compared to the exteroceptive condition (patients mean= 81.862, SD= 13.701; controls mean= 74.482, SD= 12.291). Controls had lower HR regardless of condition, but there were no interaction effects (F (1, 63)= 0.642, p= 0.426) that could be potentially confounding the significant differences found in each condition.

Regarding HRV, we found a main effect of condition: HRV was lower during interoception (F (1, 63)= 7.904, p= 0.007) in both groups. However, we did not find any group (F (1, 63)= 1.056, p= 0.308) or interaction effects (F (1, 63)= 0.619, p= 0.434) that could potentially confound our results.

Relation to symptoms

In the model using touch-related regressors to predict the BPRS total scores, the regressors STG activity and SEP spinal latency difference were slightly above significance threshold (STG: t=1.87, p=0.076; SEP: t=1.86, p=0.078).

The models including all measures (F(6,19)=1.98, p=0.142) and the interoception measures only were not significant (F(2,28)=2.48, p=0.103).

We further explored the three main subscales of the BPRS. Touch-related measures significantly predicted negative symptoms (F(4, 24) = 3.44, p = 0.027, adjusted R² = 0.289) and affective symptoms (F(4,24) = 3.115, p = 0.038, adjusted R² = 0.261). For the latter, self-related activity in the STG (t = 2.52, p = 0.02) and touch thresholds (t = -2.37, p = 0.028) contributed significantly. The touch-measures did not predict positive symptoms (F(4,24) = 0.7, p = 0.6).

Touch-interoception relationship

Neural measures of touch did not predict HEP during interoception (F(5,48) = 0.57, p = 0.72).

**Discussion**

Forward-model and social touch

Regarding the attenuation of self-produced touch sensations, it is important to mention that the forward model, that suggests a simple efference copy of the motor command to be responsible for the attenuation, appears too simple in the light of recent findings. Studies have elaborated this model and found that movement alone does not suffice attenuation^10^, indicating the need for intent for self-touch to be attenuated. Inversely, the incorrect self-attribution of touch induces attenuation even in the absence of movement^11^. The intent can be considered the sense of agency, i.e. being the cause of an action^12^. People with schizophrenia reportedly experience a decreased sense of agency^13^, particularly those with primarily negative symptoms^14^. This fits with our finding that neural measures of touch were related to negative and affective symptoms, but it contrasts with the earlier results of lowered sensory attenuation to be present mainly in those with positive symptomatology^15^.

For other-touch, patients showed increased activity in a temporoparietal cluster that overlaps with the secondary somatosensory cortex, posterior insula, and temporo-parietal junction. This region of interest was functionally defined on our previous findings on self-other-touch processing changes during a pharmacological manipulation of the sense of self using ketamine^16^. We found a reduction of other-related activity in this area during the ketamine session – the opposite from our here reported findings in schizophrenia. This is especially of interest because ketamine has been suggested as a pharmacological model for schizophrenia before^17,18^. A potential explanation for this disparity could be that ketamine can be understood as a model for acute or early stage psychosis, while our patients had been diagnosed for several years and were medicated. However, it is also possible that our findings indicate a different underlying mechanism in schizophrenia than during ketamine administration. Finally, a more basic explanation could be that schizophrenia patients experience less social affective touch in their daily life than controls^19^ and it has been shown that touch deprivation relates to social touch perception^20^. Therefore, an increased neural response to social touch in schizophrenia might simply indicate that this stimulus is more salient, relevant, or surprising. We did however not find increased responses to social touch in its primary processing areas, the primary somatosensory cortex and the insula.

**Tables**:

**Table S1. Sample who completed the heartbeat tapping task**

| **Basic Demographics** | Control | Patient | Test variable | Significance |
| --- | --- | --- | --- | --- |
| Gender | 15F/19M | 14F/20M | x^2^ (1) = 0.060, | p = 0.806 |
| Age | 35.5 (9.4) | 35.6 (7.5) | t (66) = -0.043, | p = 0.966 |
| BMI | 24.3 (4.3) | 29.4 (6.0) | t (66) = -4.039, | p < 0.001** |
| **Handedness** |  |  | x^2^ (2) = 4.410 | p = 0.110 |
| Right | 33 (97.1%) | 28 (82.4%) |  |  |
| Left | 0 (0%) | 3 (8.8%) |  |  |
| Ambidextrous | 1 (2.9%) | 3 (8.8%) |  |  |
| **Education** |  |  | x^2^ (2) = 7.111 | p = 0.029* |
| Primary | 0 (0%) | 5 (14.7%) |  |  |
| Secondary | 12 (35.3%) | 15 (44.1%) |  |  |
| Tertiary | 22 (64.7%) | 14 (41.2%) |  |  |
| **Occupation** |  |  | x^2^ (2) = 24.571 | p < 0.001** |
| Unemployed / sick | 0 (0%) | 4 (11.8%) |  |  |
| Student | 10 (29.4%) | 18 (52.9%) |  |  |
| Working | 24 (70.6%) | 12 (35.3%) |  |  |
| **Questionnaires** |  |  |  |  |
| MAIA | 101.7 (21.8) | 87.6 (18.6) | t (66) = 2.872 | p = 0.005** |
| MAIA: Noticing | 3.6 (0.9) | 3.2 (0.9) | t (66) = 2.068 | p = 0.043* |
| MAIA: Non-distracting | 2.5 (0.8) | 2.1 (0.8) | t (66) = 1.823 | p = 0.073 |
| MAIA: Non-worrying | 3.1 (0.9) | 2.3 (1.2) | t (66) = 3.098 | p = 0.003** |
| MAIA: Attention regulation | 3.2 (0.9) | 2.9 (0.9) | t (66) = 1.449 | p = 0.152 |
| MAIA: Emotional awareness | 3.4 (0.9) | 3.0 (1.0) | t (66) = 1.721 | p = 0.090 |
| MAIA: Self-regulation | 3.0 (1.1) | 2.5 (1.1) | t (66) = 1.851 | p = 0.069 |
| MAIA: Bodily listening | 2.5 (1.1) | 2.5 (1.1) | t (66) > 0.001 | p = 1.000 |
| MAIA: Trusting | 3.8 (1.0) | 3.0 (1.4) | U = 783.5, | p = 0.011* |
| STQ | 27.3 (9.2) | 34.0 (14.2) | U = 429.5 | p = 0.069 |
| EQ | 46.3 (13.1) | 41.3 (13.0) | t (66) = 1.590, | p = 0.117 |
| SP: Low Registration | 28.6 (5.6) | 34.0 (7.6) | U = 322.0 | p = 0.002** |
| SP: Sensation Seeking | 42.7 (5.9) | 41.2 (7.6) | t (66) = 0.928, | p = 0.357 |
| SP: Sensation Sensitivity | 33.4 (5.4) | 37.7 (9.5) | U = 402.5, | p = 0.032* |
| SP: Sensation Avoiding | 36.3 (7.0) | 40.9 (9.2) | t (66) = -2.296, | p = 0.025* |
| SP: Touch processing | 29.1 (3.8) | 30.5 (6.7) | U = 495.0, | p = 0.310 |
| *Note. *Significant but does not survive correction for multiple comparisons. **Significant after correction.* | | | | |

**Table S2. Sample who completed the somatosensory evoked potentials**

| **Basic Demographics** | Control | Patient | Test variable | Significance |
| --- | --- | --- | --- | --- |
| Gender | 15F/20M | 14F/20M | x^2^ (1) = 0.060, | p = 0.806 |
| Age | 35.7 (9.3) | 35.5 (7.5) | t (66) = -0.043, | p = 0.966 |
| BMI | 24.3 (4.3) | 29 (6.2) | t (66) = -4.039, | p < 0.001** |
| **Handedness** |  |  | x^2^ (2) = 4.410 | p = 0.110 |
| Right | 34 (97.1%) | 28 (82.4%) |  |  |
| Left | 0 (0%) | 3 (8.8%) |  |  |
| Ambidextrous | 1 (2.9%) | 3 (8.8%) |  |  |
| **Education** |  |  | x^2^ (2) = 7.802, | p = 0.020* |
| Primary | 0 (0%) | 6 (17.6%) |  |  |
| Secondary | 13 (37.1%) | 14 (41.2%) |  |  |
| Tertiary | 22 (65.9%) | 14 (41.2%) |  |  |
| **Occupation** |  |  | x^2^ (2) = 23.225 | p < 0.001* |
| Unemployed / sick | 0 (0%) | 17 (50.0%) |  |  |
| Student | 10 (28.6%) | 5 (14.7%) |  |  |
| Working | 25 (71.4%) | 12 (35.3%) |  |  |
| **Questionnaires** |  |  |  |  |
| MAIA | 101.5 (21.5) | 87.2 (18.6) | t (67) = 2.958 | p = 0.004** |
| MAIA: Noticing | 3.6 (0.8) | 3.2 (0.9) | t (67) = 2.012 | p = 0.048* |
| MAIA: Non-distracting | 2.4 (0.8) | 2.1 (0.8) | t (67) = 1.854 | p = 0.068 |
| MAIA: Non-worrying | 3.1 (0.9) | 2.3 (1.2) | t (67) = 3.136 | p = 0.003** |
| MAIA: Attention regulation | 3.2 (0.9) | 2.9 (1.0) | t (67) = 1.199 | p = 0.235 |
| MAIA: Emotional awareness | 3.4 (0.9) | 2.9 (1.1) | t (67) = 2.057 | p = 0.044* |
| MAIA: Self-regulation | 3.0 (1.0) | 2.4 (1.1) | t (67) = 2.029 | p = 0.046* |
| MAIA: Bodily listening | 2.5 (1.1) | 2.5 (1.2) | t (67) = 0.157 | p = 0.876 |
| MAIA: Trusting | 3.8 (1.0) | 3.0 (1.4) | U = 790.0 | p = 0.018* |
| STQ | 37.2 (9.1) | 44.4 (14.1) | U = 419.0 | p = 0.035* |
| EQ | 46.1 (13.0) | 41.7 (12.9) | t (67) = 1.413 | p = 0.162 |
| SP: Low Registration | 28.5 (5.6) | 33.9 (7.6) | U = 330.5 | p = 0.001** |
| SP: Sensation Seeking | 42.6 ( 5.9) | 40.9 (7.5) | t (67) = 1.044 | p = 0.300 |
| SP: Sensation Sensitivity | 33.1 (5.6) | 37.9 (9.7) | U = 399.0 | p = 0.019* |
| SP: Sensation Avoiding | 36.1 (7.1) | 41.1 (9.0) | t (67) = -2.581 | p = 0.012* |
| SP: Touch processing | 28.9 (4.1) | 30.4 (6.7) | U = 505.0 | p = 0.282 |
| *Note. *Significant but does not survive correction for multiple comparisons using Bonferroni-Holms. **Significant after correction.* | | | | |

**Table S3. Sample who completed the touch threshold task.**

| **Basic Demographics** | Control | Patient | Test variable | Significance |
| --- | --- | --- | --- | --- |
| Gender | 15F/20M | 14F/20M | x^2^ (1) = 0.035 | p = 0.851 |
| Age | 35.7 (9.3) | 35.7 (7.3) | U = 550.5 | p = 0.923 |
| BMI | 24.3 (4.3) | 29 (6.2) | t (64) = -3.846 | p < 0.001** |
| **Handedness** |  |  | x^2^ (2) = 3.838 | p = 0.147 |
| Right | 34 (97.1%) | 26 (83.9%) |  |  |
| Left | 0 (0%) | 2 (6.4%) |  |  |
| Ambidextrous | 1 (2.9%) | 3 (9.7%) |  |  |
| **Education** |  |  | x^2^ (2) = 8.731 | p = 0.013** |
| Primary | 0 (0%) | 6 (19.4%) |  |  |
| Secondary | 13 (37.1%) | 13 (41.9%) |  |  |
| Tertiary | 22 (62.9%) | 12 (38.7%) |  |  |
| **Occupation** |  |  | x^2^ (2) = 21.949 | p < 0.001** |
| Unemployed / sick | 0 (0%) | 15 (48.4%) |  |  |
| Student | 10 (28.6%) | 5 (16.1%) |  |  |
| Working | 25 (71.4%) | 11 (35.5%) |  |  |
| **Questionnaires** |  |  |  |  |
| MAIA | 101.5 (21.5) | 85.8 (18.6) | t (64) = 3.167 | p = 0.002** |
| MAIA: Noticing | 3.6 (0.8) | 3.2 (0.9) | t (64) = 1.926 | p = 0.059 |
| MAIA: Non-distracting | 2.4 (0.8) | 2.1 (0.8) | t (64) = 1.768 | p = 0.082 |
| MAIA: Non-worrying | 3.1 (0.9) | 2.4 (1.2) | t (64) = 2.735 | p = 0.008* |
| MAIA: Attention regulation | 3.2 (0.9) | 2.9 (1.0) | t (64) = 1.250 | p = 0.216 |
| MAIA: Emotional awareness | 3.4 (0.9) | 2.9 (1.2) | t (64) = 2.108 | p = 0.039* |
| MAIA: Self-regulation | 3.0 (1.0) | 2.3 (1.0) | t (64) = 2.594 | p = 0.012* |
| MAIA: Bodily listening | 2.5 (1.1) | 2.3 (1.1) | t (64) = 0.737 | p = 0.464 |
| MAIA: Trusting | 3.8 (1.0) | 2.9 (1.3) | U = 753.5 | p = 0.006** |
| STQ | 37.2 (9.1) | 45.0 (14.4) | U = 371.0 | p = 0.028* |
| EQ | 46.1 (13.0) | 41.4 (12.8) | t (64) = 1.467 | p = 0.147 |
| SP: Low Registration | 28.5 (5.6) | 34.6 (7.3) | U = 276.0 | p < 0.001** |
| SP: Sensation Seeking | 42.6 (5.9) | 41.1 (7.7) | t (64) = 0.864 | p = 0.391 |
| SP: Sensation Sensitivity | 33.1 (5.6) | 38.9 (9.3) | U = 322.5 | p = 0.005** |
| SP: Sensation Avoiding | 36.1 (7.1) | 41.3 (9.4) | t (64) = -2.589 | p = 0.012 |
| SP: Touch processing | 28.6 (4.1) | 31.2 (6.1) | U = 422.5 | p = 0.124 |
| *Note. *Significant but does not survive correction for multiple comparisons using Bonferroni-Holms. **Significant after correction.* | | | | |

**Table S4. Sample who completed the functional magnetic resonance imaging.**

| **Basic Demographics** | Control | Patient | Test variable | Significance |
| --- | --- | --- | --- | --- |
| Gender | 11F/20M | 11F/17M | x^2^ (1) = 0.091 | p = 0.763 |
| Age | 34.8 (9.1) | 35.5 (7.7) | t (57) = -0.264 | p = 0.793 |
| BMI | 24.0 (4.1) | 28.9 (6.0) | t (57) = -3.680 | p < 0.001** |
| **Handedness** |  |  | x^2^ (2) = 5.091 | p = 0.078 |
| Right | 30 (96.7%) | 22 (78.6%) |  |  |
| Left | 0 (0%) | 3 (10.7%) |  |  |
| Ambidextrous | 1 (3.3%) | 3 (10.7%) |  |  |
| **Education** |  |  | x^2^ (2) = 8.776 | p = 0.012** |
| Primary | 0 (0%) | 6 (20.7%) |  |  |
| Secondary | 12 (38.7%) | 13 (44.8%) |  |  |
| Tertiary | 19 (61.3%) | 10 (34.5%) |  |  |
| **Occupation** |  |  | x^2^ (2) = 19.652 | p < 0.001** |
| Unemployed / sick | 0 (0%) | 14 (48.3%) |  |  |
| Student | 10 (32.3%) | 4 (13.8%) |  |  |
| Working | 21 (67.7%) | 11 (37.9%) |  |  |
| **Questionnaires** |  |  |  |  |
| MAIA | 99.3 (21.4) | 88.3 (19.1) | t (57) = 2.088 | p = 0.041* |
| MAIA: Noticing | 3.6 (0.8) | 3.3 (0.8) | t (57) = 1.458 | p = 0.150 |
| MAIA: Non-distracting | 2.4 (0.8) | 1.9 (0.8) | t (57) = 2.079 | p = 0.042* |
| MAIA: Non-worrying | 3.1 (0.9) | 2.4 (1.3) | t (57) =2.322 | p = 0.024* |
| MAIA: Attention regulation | 3.1 (0.9) | 3.0 (0.9) | t (57) = 0.667 | p = 0.508 |
| MAIA: Emotional awareness | 3.4 (0.9) | 3.0 (1.1) | t (57) = 1.459 | p = 0.150 |
| MAIA: Self-regulation | 2.9 (1.1) | 2.4 (1.2) | t (57) = 1.672 | p = 0.100 |
| MAIA: Bodily listening | 2.4 (1.0) | 2.5 (1.3) | t (57) = -0.529 | p = 0.599 |
| MAIA: Trusting | 3.8 (1.0) | 3.1 (1.3) | U = 546.0 | p = 0.087 |
| STQ | 37.6 (9.3) | 44.6 (14.0) | U = 314.0 | p = 0.069 |
| EQ | 45.5 (13.5) | 40.7 (12.7) | t (57) = 1.397 | p = 0.168 |
| SP: Low Registration | 28.3 (5.6) | 34.6 (7.2) | t (57) = -3.766 | p < 0.001** |
| SP: Sensation Seeking | 42.3 (5.3) | 40.7 (7.8) | t (57) = 0.923 | p = 0.360 |
| SP: Sensation Sensitivity | 33.0 (5.7) | 38.2 (9.5) | U = 269.5 | p = 0.013* |
| SP: Sensation Avoiding | 36.0 (7.2) | 41.6 (9.5) | t (57) = -2.560 | p = 0.013* |
| SP: Touch processing | 29.2 (4.1) | 30.1 (6.8) | U = 401.0 | p = 0.069 |
| *Note. *Significant but does not survive correction for multiple comparisons using Bonferroni-Holms. **Significant after correction.* | | | | |

**Table S5: Main effect of group.**

| **Patients > Controls** |  |  |  |  |  |  |  |
| --- | --- | --- | --- | --- | --- | --- | --- |
| **Region** | ***k*** | **L/R** | **x** | **y** | **z** | ***t*** | ***p(FWE)*** |
| Secondary Visual cortex | 564 | R | 16 | -92 | 12 | 14.87 | <0.001 |
|  |  |  | 26 | -86 | 14 | 11.61 | <0.001 |
|  |  |  | 38 | -84 | 4 | 9.22 | <0.001 |
|  | 23 | L | -24 | -88 | 2 | 6.78 | <0.001 |
| Medial temporal gyrus | 507 | R | 54 | -50 | 6 | 8.98 | <0.001 |
|  |  |  | 42 | -66 | 8 | 7.90 | <0.001 |
|  |  |  | 52 | -64 | 2 | 5.70 | <0.001 |
| Fusiform gyri | 345 | R | 28 | -66 | -12 | 8.73 | <0.001 |
|  |  |  | 30 | -78 | -16 | 7.95 | <0.001 |
|  |  |  | 26 | -82 | -10 | 7.52 | <0.001 |
| Primary Somatosensory cortex | 1058 | R | 50 | -20 | 40 | 8.33 | <0.001 |
|  |  |  | 48 | -28 | 12 | 8.25 | <0.001 |
|  |  |  | 60 | -44 | 26 | 8.07 | <0.001 |
|  | 31 | R | 32 | -46 | 66 | 7.15 | <0.001 |
|  | 86 | L | -10 | -34 | 70 | 6.95 | <0.001 |
|  |  |  | 0 | -36 | 64 | 5.94 | 0.001 |
|  |  |  | -10 | -44 | 72 | 5.39 | 0.010 |
| Premotor cortex | 162 | R | 44 | -8 | 56 | 7.90 | <0.001 |
|  |  |  | 38 | -10 | 64 | 7.35 | <0.001 |
|  | 30 | R | 52 | 2 | 32 | 6.41 | <0.001 |
|  |  |  | 36 | -18 | 66 | 7.33 | <0.001 |
| Angular gyrus | 441 | R | -54 | -44 | 26 | 7.77 | <0.001 |
|  |  |  | -46 | -30 | 10 | 7.09 | <0.001 |
|  |  |  | -54 | -32 | 14 | 7.02 | <0.001 |
| Supramarginal gyrus | 114 | R | 36 | -40 | 52 | 7.65 | <0.001 |
|  |  |  | 42 | -36 | 62 | 6.01 | 0.001 |
|  |  |  | 44 | -32 | 54 | 5.29 | 0.014 |
|  | 69 | L | -52 | -30 | 44 | 7.17 | <0.001 |
|  |  |  | -56 | -36 | 48 | 5.77 | 0.002 |
| Secondary Somatosensory cortex | 49 | R | 10 | -38 | 58 | 7.44 | <0.001 |
| Dorsal Posterior cingulate cortex | 454 | R | 4 | -26 | 46 | 7.21 | <0.001 |
|  |  |  | 2 | -12 | 42 | 6.57 | <0.001 |
|  |  |  | 2 | -8 | 34 | 6.54 | <0.001 |
| Visual cortex | 119 | L | -46 | -66 | 10 | 7.13 | <0.001 |
| Operculum | 77 | R | 54 | 4 | 0 | 6.50 | <0.001 |
| Primary Motor cortex | 40 | R | 56 | -2 | 16 | 6.37 | <0.001 |
| Insula | 35 | R | 40 | -10 | -14 | 6.17 | <0.001 |

| **Controls > Patients** |  |  |  |  |  |  |  |
| --- | --- | --- | --- | --- | --- | --- | --- |
| **Region** | **k** | **L/R** | **x** | **y** | **z** | **t** | **p(FWE)** |
| Secondary visual cortex | 99 | R | 12 | -80 | -2 | 12.72 | <0.001 |
|  | 517 | L | -12 | -82 | -12 | 10.55 | <0.001 |
|  |  |  | -10 | -78 | 4 | 8.39 | <0.001 |
|  |  |  | -10 | -94 | 2 | 8.36 | <0.001 |
| Calcarine cortex | 148 | R | 6 | -86 | 8 | 9.13 | <0.001 |
|  |  |  | 16 | -80 | 10 | 7.39 | <0.001 |
| Cuneus | 26 | R | 10 | -90 | 22 | 7.16 | <0.001 |

**Table S6.: Logistic regression analysis of neural measures on group membership**

| Independent variable | Coefficient estimate | Standard error | Z | P | Odds Ratio |
| --- | --- | --- | --- | --- | --- |
| Intercept | -0.431 | 0.802 | -0.538 | 0.591 | 0.65 |
| STG self | 3.967 | 1.458 | 2.721 | 0.007 | 52.803 |
| TPC other | 1.135 | 1.006 | 1.129 | 0.259 | 3.112 |
| Cerv.N13 self-other | -0.836 | 0.507 | -1.649 | 0.099 | 0.433 |
| HEP Intero | 0.048 | 0.0032 | 1.488 | 0.137 | 1.049 |

**Figure S1: Flow-chart of participant attrition.** HBD = heartbeat detection task, SEP = somatosensory evoked potentials, TT = touch thresholds, fMRI = functional magnetic resonance imaging
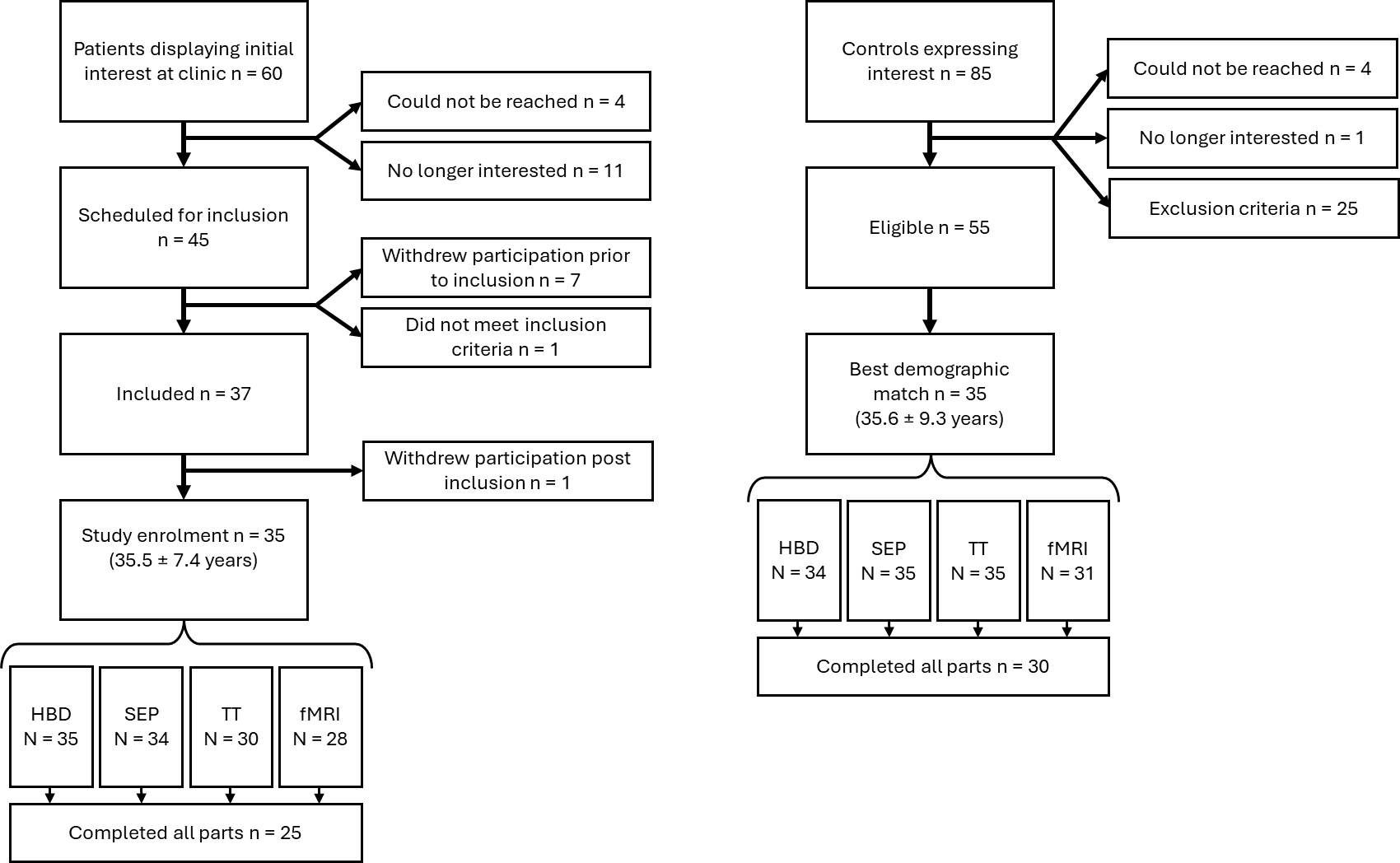

**Figure S2: Patients show increased brain activity compared to the controls across touch conditions (main effect of group).** Thresholded at p<0.001 for display purpose, color-bar indicates t-values.

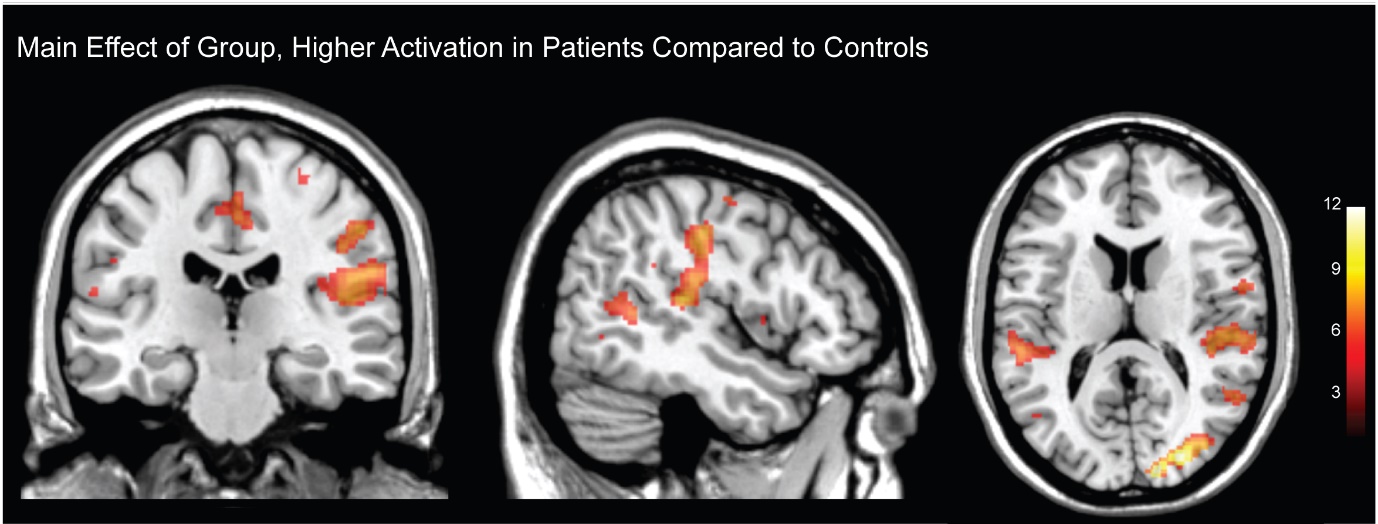
